# Sharing Aggregated Patient Counts in Place of Line-Level EHR Data: Analytic Fidelity and the Limits of Count Suppression for Privacy

**DOI:** 10.64898/2026.08.18.26359984

**Authors:** Yanshi Chen, Andrew J. McMurry, Daniel Gottlieb, James R. Jones, Benjamin J. Strober, Kenneth D. Mandl

**Affiliations:** Computational Health Informatics Program, Boston Children’s Hospital, Boston, MA, USA; Department of Biomedical Informatics, Harvard Medical School, Boston, MA, USA; Department of Pediatrics, Harvard Medical School, Boston, MA, USA; Central Square Solutions, Boston, MA, USA

**Keywords:** electronic health records, data privacy, statistical data analysis, kidney transplantation, data sharing, probability, Bayes’ theorem, cohort studies

## Abstract

**Objective:** Privacy regulation constrains sharing line-level electronic health records (EHR) across institutions. One alternative is to aggregate counts into a cube, a table of counts for every combination of categorical variables, with cells below a threshold suppressed. This study asked whether common analyses on the cube reproduce conclusions from line-level data, and whether suppression prevents recovery of the small cells it is meant to hide.

**Materials and Methods:** A Bayesian count-inference pipeline was built that reconstructs suppressed counts and doubles as a reconstruction attack. Applied to 285 pediatric kidney-transplant patients at Boston Children’s Hospital, statistical fidelity (Jensen-Shannon divergence, Cramér’s V, and R²) and analytical utility (marginal distributions, subgroup graft rejection odds ratios, and logistic-regression classification) were evaluated. Conditional Tabular GAN (CTGAN) synthetic data served as a comparator.

**Results:** Statistical analyses on the cube recapitulated results from line-level data. Across 106 demographic-by-medication subgroups, a bootstrap mean of 3.5 subgroups showed a significant graft-rejection association. The cube’s odds-ratio sign changes reversed no significant associations, versus 2.3 for CTGAN. The same reconstruction also defeated suppression: in a 10-variable cube, 76.6% of suppressed cube cells were recovered exactly (14,554 of 18,994), including 85.5% of single-patient cells.

**Discussion:** The cube reproduced common kidney-transplant analyses, but the same reconstruction also recovered suppressed cells; fidelity and privacy risk are thus two faces of one reconstruction rather than independent properties.

**Conclusions:** The cube is a useful surrogate for these kidney-transplant analyses only when paired with a stronger privacy mechanism. This study demonstrated reconstructability of suppressed counts, not re-identification.

## Background and Significance

Electronic health records (EHR) support patient care, public health, research, and the development of clinical AI,[1, 2] yet realizing much of that value requires pooling data across institutions for multi-site retrospective cohort studies, and sharing line-level records between sites is slow, costly, and constrained by privacy law such as HIPAA. One alternative to exchanging identifiable records is to release only aggregated patient counts, suppressing any cell below a small threshold so that no individual can be singled out. This study examines one such release format, a cube of patient counts, the union of cross-tabulations over every subset of the grouping variables, and asks two questions of it: can analyses run on the cube reproduce what the same analyses would have found on line-level data, and can the suppressed counts be recovered, undoing the protection they are meant to provide?

Specifically, we (1) develop a Bayesian count-inference pipeline that infers the suppressed cube cells; (2) evaluate statistical fidelity of the cube using distributional, association, and reconstruction metrics; (3) evaluate analytical utility of the cube through descriptive, inferential, and predictive tasks typical in transplant research; and (4) evaluate the privacy preservation the cube provides. These questions are evaluated in the context of pediatric kidney transplantation, a clinically important use case in which immunosuppressive regimens must balance graft rejection against infection,[3, 4] individual responses to therapy vary substantially,[5] and outpatient EHR-based outcomes analysis has precedent.[6]

## Materials and Methods

The methods inferred the counts hidden by suppression, then evaluated the statistical fidelity and analytical utility of the resulting inferred cube against the original line-level data. The cube and Conditional Tabular GAN (CTGAN) were compared as two alternatives to sharing line-level records, the cube suppressing small counts and CTGAN releasing synthetic patient data.

### Data

The study cohort comprised pediatric patients receiving care for a kidney transplant at Boston Children’s Hospital between January 1, 2021 and December 31, 2023. Expert-curated diagnoses and procedure codes for kidney transplant-related care events were used, and at least two clinical encounters during the study period were required. The final cohort contained 285 patients, from whom demographics, conditions, and medications were extracted. The BCH Committee on Clinical Investigation (BCH IRB-P00043392) determined the study to be exempt from full human participant oversight. A waiver of consent was obtained to allow corpus extraction and chart review for data quality purposes.

### Study variables

The study variables comprised demographics, immunosuppressive medication classes, and graft rejection. Three demographic variables were used: age, race, and sex. Age was discretized into one bin per year over 0–21 years (all patients were ≤21 years old), yielding 22 categories. Race had six categories: White, Black or African American, Asian, American Indian or Alaska Native, Other race, and Asked but unknown. Sex was defined as male or female.

Immunosuppressive exposure was represented by seven medication classes: anti-metabolite, calcineurin inhibitor (CNI), corticosteroid, costimulation blocker, monoclonal antibody (mAb), polyclonal antibody, and mTOR inhibitor. Medications were matched to RxNorm ingredients and grouped into these classes using a curated value set of ingredient synonyms spanning brand, generic, ingredient, and formulation names; the full coded value set is provided in Supplementary Table S1. Each class was encoded as an “ever exposed” binary indicator. Graft rejection was encoded as a single binary variable, coded positive when the patient’s record contained any diagnosis in the Cumulus Library kidney-transplant graft-rejection value set, a set of 23 ICD-10-CM and SNOMED CT codes spanning hyperacute, acute, and chronic rejection through unspecified transplant complications.[7]

### Cube construction

A cube was generated as the output of a Trino cube operation[8, 9]: for a dataset of N records described by v categorical variables, it returns aggregate counts for every element of the power set of grouping variables. The empty grouping yields the grand total N; single-variable groupings give marginal counts; two-variable groupings yield pairwise cross-tabulations; and grouping by all v variables yields the most granular, atomic cells, each a unique combination of variable levels and therefore a distinguishable group of patients. Any count below k=10 was suppressed (replaced with null) before release. Two cubes were analyzed: race x sex x age (3 variables, 264 cells), whose high-cardinality age variable (one category per year) left atomic cells sparse; and race x sex x seven binary immunosuppressive medication classes x graft rejection (10 variables, 25,488 cells), combining higher dimensionality with mostly binary, low-cardinality variables. The two differ simultaneously in dimensionality, cardinality, and atomic-cell sparsity.

### Count inference

The suppressed counts were inferred with a four-stage pipeline combining exact recovery with Bayesian estimation: (1) translate the released cube into a constraint system by writing each aggregate as the sum of its constituent atomic cells, so each published aggregate gives a linear equality and each suppressed aggregate a bounded inequality; (2) solve two integer linear programs (ILP) per atomic cell to bound its count, deterministically recovering every cell whose lower and upper bounds coincide; (3) for the remaining cells, fit a Bayesian Poisson-log-normal model to the published and suppressed counts and sample the atomic-cell rates; (4) conditional on each rate sample, draw integer count vectors consistent with all constraints and take the posterior mean and re-aggregate to cube format as a single point-estimate reconstruction. Every published aggregate establishes a linear equation: the sum of its constituent atomic-cell counts equals the published value. Every suppressed aggregate establishes an inequality: its constituent sum lies in [0, k). Following Abowd et al.[10] and Dobra and Fienberg,[11] we used two integer linear programs (ILP) per atomic cell, one minimizing and one maximizing the cell’s count subject to all published equalities and inequalities, to obtain the tightest feasible range. When the lower and upper bounds coincide, the suppressed count is deterministically recovered. For cells whose bounds do not coincide, a Bayesian model inferred the most likely value from a prior over atomic-cell rates (each cell’s expected count), yielding a posterior distribution over feasible counts.[12] Each atomic cell *j* was modeled as an independent draw from a Poisson distribution with rate parameter *λ_j_*, its expected count. A log-normal prior on the rate parameters *λ_j_* ensured positive rates while remaining weakly informative across the range of atomic-cell counts in the sparse cube:

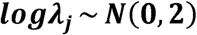

Each aggregate count follows a Poisson distribution with a rate equal to the sum of rates of its constituent atomic cells. The model incorporated two types of observed data:

For published aggregate count *y_i_* over a subset *S_i_* of atomic cells, the observed count is Poisson distributed with rate equal to the sum of the rates of its constituent cells.

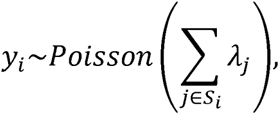

For each suppressed count over a subset *S_i_* of atomic cells, the count is known to be less than the suppression threshold *k*. The likelihood contribution is the probability that a Poisson random variable with rate equal to the sum of the constituent cell rates has a value below the suppression threshold *k*:

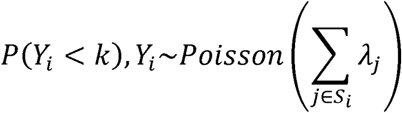

Posterior rates were sampled with the No-U-Turn Sampler (2 chains, 100 warmup and 200 sampling iterations per chain, yielding 400 posterior rate draws).[13] Conditional on each rate sample, integer atomic-cell counts that exactly satisfied all published equalities and ILP-derived bounds were drawn by Metropolis–Hastings (2000 iterations thinned by 10).[14] The posterior mean of the atomic-cell count vectors served as the point estimate for each suppressed cell and was re-aggregated to cube format as a single reconstruction for all fidelity and utility analyses.

### Baseline comparator

The cube’s fidelity and utility were judged against a comparator that releases something other than line-level data. Synthetic data, in which a generative model trained on the original records produces new pseudo-records, is the most widely used such alternative for tabular EHR.[15, 16] CTGAN[17], a standard benchmark for tabular synthetic data, was applied to each analysis alongside the cube. Differences between the cube and CTGAN on the same analysis indicate how much of a conclusion depends on the release method rather than the underlying data. Baselines for the inference task included (i) imputing all suppressed counts as 0, (ii) as the suppression midpoint of 5, (iii) as uniformly random integers in [0, 9], and (iv) treating CTGAN synthetic line-level data as a source of inferred counts. Inference accuracy was scored as the mean absolute error (MAE) between each method’s estimates and the true counts, computed over the atomic cells suppressed in the released cube. CTGAN was implemented with the SDV CTGANSynthesizer, metadata auto-detected from the cohort, fit on the line-level data with default hyperparameters, and sampled to produce a synthetic dataset of equal size. We treat CTGAN not as a tuned, competitive benchmark but as an off-the-shelf, untuned comparator, trained without hyperparameter optimization on a small cohort.

### Fidelity evaluation

Statistical fidelity was assessed using three complementary measures, each computed between the original line-level data and either the inferred cube (posterior-mean count) or the CTGAN sample (a single synthetic draw matching the cohort size). Jensen–Shannon divergence (JSD)[18], bounded in [0, 1] with 0 indicating identical distributions, was computed for every marginal and for the full joint distribution. Cramér’s V (a 0-to-1 measure of association between two categorical variables)[19] was computed for every variable pair; preservation was assessed by whether the cube’s or CTGAN’s value fell within the original’s confidence interval, and summarized by the mean absolute difference across all pairs. The coefficient of determination (R²) was computed between the original and the inferred cube atomic-cell count vectors.

Uncertainty was quantified identically for every measure using nonparametric bootstrapping with 2,000 replicates: the original cohort was resampled with replacement and evaluation statistics were computed against the inferred cube (posterior-mean) and the CTGAN sample each time. Each statistic was reported as its bootstrap-mean point estimate with a 95% confidence interval (CI).

### Utility evaluation

Three downstream analyses were chosen to mirror common transplant-research workflows. First, marginal probability distributions P(sex), P(race), and P(age), including the cumulative distribution of age, were estimated and compared to the originals. Second, graft-rejection odds ratios were computed for every subgroup in the race x sex x 7 medication class x graft rejection cube, comparing the odds of rejection within each subgroup against the union of the remaining subgroups. We tracked both the absolute difference in log odds ratios and any sign changes (subgroups whose direction of association would be reported differently). A sign change is clinically consequential if it reverses a statistically significant association, so each flipped subgroup was tested by Fisher’s exact test on its subgroup-vs-rest x graft rejection table, and flips reaching a nominal p < 0.05 were counted, with significance assessed on the resampled original data under the same bootstrap. Third, a multivariate logistic-regression classifier was trained to predict graft rejection from race, sex, and the seven medication-class indicators. The original data were split 80/20 for training and testing with stratification, respectively. On the training set, we formed the posterior-mean cube reconstruction and a CTGAN sample and trained one classifier on each source. A classifier was also trained on the original line-level training data. All three were evaluated on the test set by accuracy, precision, recall, and F1, with 95% CI from nonparametric bootstrapping across 2000 test-set resamples.

## Results

Across every evaluation, the inferred cube reflected the line-level data more closely than the CTGAN comparator, with high statistical fidelity on all three measures that carried through to the descriptive, inferential, and predictive analyses.

### Count inference recovers many suppressed counts in the more densely constrained Cube

The race x sex x age cube produced 264 cells, of which 50 were published directly; of the 214 suppressed cells, 12 (5.6%) were deterministically recovered by ILP and 202 (94.4%) were probabilistically inferred. The race x sex x 7 medication class x graft rejection cube produced 25,488 cells, 6,494 published directly and 18,994 suppressed; of the suppressed cells, 14,554 (76.6%) were deterministically recovered and 4,440 (23.4%) were probabilistically inferred. The deterministic-recovery rate among very small atomic cells in this cube was substantial: 47 of 55 single-patient cells (85.5%), 13 of 18 two-patient cells (72.2%), and 5 of 7 three-patient cells (71.4%) could be exactly recovered from the published counts alone.

Mean absolute error (MAE) of each method’s suppressed-cell estimates against the true counts, with 95% CI from the bootstrap, is summarized in Table 1. In the 3-variable cube, the inference pipeline achieved the lowest MAE (1.64; 95% CI, 1.43–1.86), outperforming the synthetic data approach (2.06, 95% CI, 1.83-2.29) and the other baseline methods. In the 10-variable cube, MAE dropped to 1.06 (95% CI, 0.89–1.25), outperforming the synthetic data approach (1.79, 95% CI, 1.56-2.02) and the other baseline methods. The stronger recovery in the 10-variable cube reflects its denser, more overlapping system of published aggregates (see Discussion).

**Table 1.** Mean absolute error (MAE) of suppressed-count estimates against true counts, by cube and baseline methods.

| **Method** | **race x sex x age MAE (95% CI)** | **race x sex x 7 med class x rejection MAE (95% CI)** |
| --- | --- | --- |
| **Count Inference** | 1.64 (1.43, 1.86) | 1.06 (0.89, 1.25) |
| **Suppressed = 0** | 2.58 (2.49, 2.65) | 2.10 (1.95, 2.26) |
| **Suppressed = 5** | 3.12 (2.88, 3.38) | 3.37 (3.15, 3.60) |
| **Suppressed = random (0-9)** | 3.50 (3.23, 3.77) | 3.80 (3.55, 4.04) |
| **CTGAN synthetic data** | 2.06 (1.83, 2.29) | 1.79 (1.56, 2.02) |

### Cube-derived data preserves marginal and joint distributions

For each feature, the marginal Jensen–Shannon divergence (JSD) of the inferred cube and of CTGAN relative to the line-level data is shown in Figure 1. In both cubes, the inferred cube’s JSD and its interval lay at or below CTGAN’s. In the race x sex x age cube, marginal JSD against the original was 0.0004 (95% CI, 0-0.0021) for sex, 0.008 (0.002-0.015) for race, and 0.013 (0.007-0.022) for age, compared with 0.002 (0-0.006), 0.02 (0.01-0.03), 0.07 (0.05-0.09) for CTGAN. JSD for the full joint distribution was 0.102 (0.081-0.124) for the cube versus 0.273 (0.247-0.300) for CTGAN. In the race x sex x 7 medication class x graft rejection cube, all marginal JSDs were near zero; the joint JSD was 0.059 (0.046–0.073) versus 0.218 (0.194-0.244) for CTGAN.

**Figure 1.**
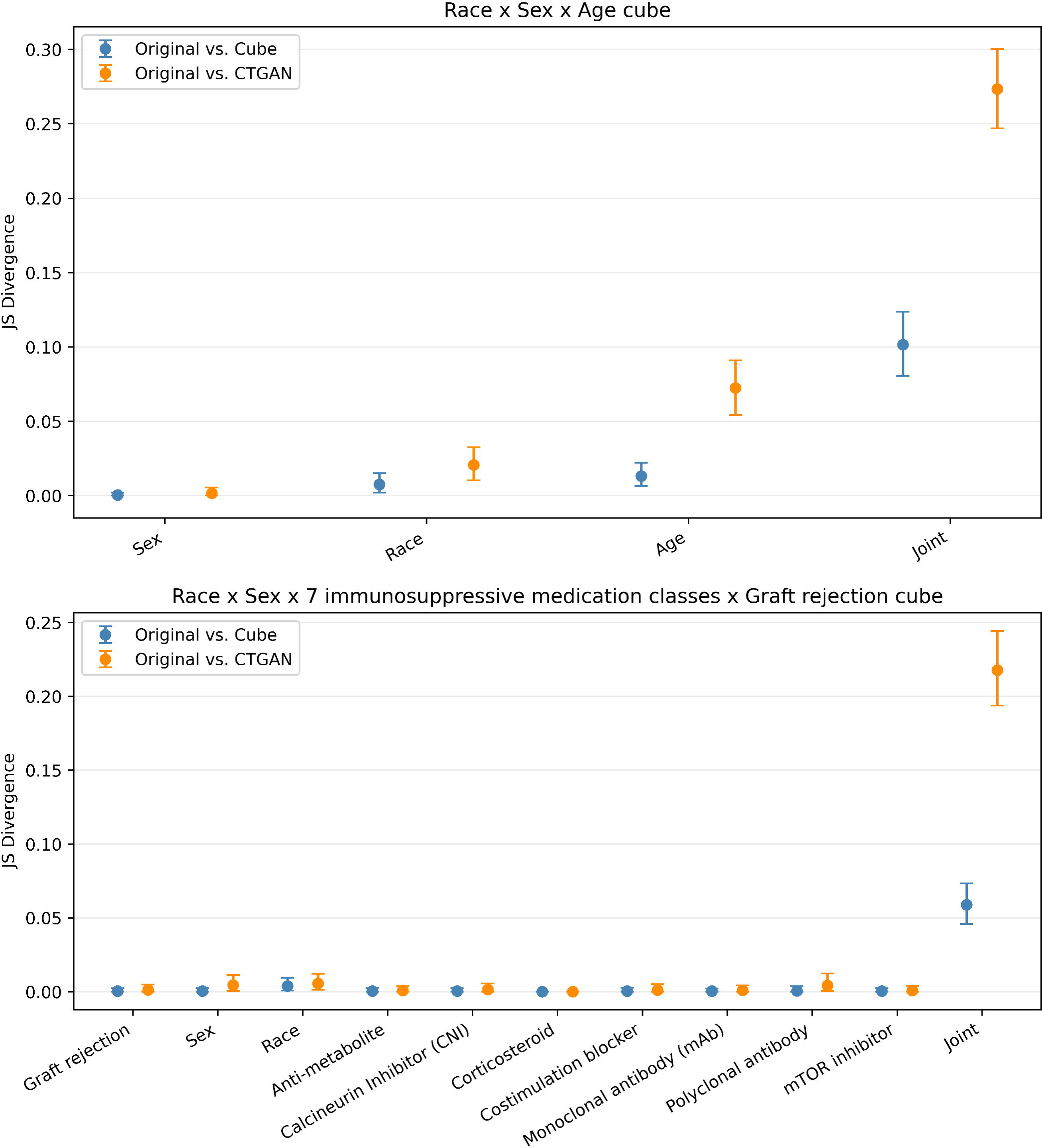
Jensen–Shannon divergence against the original data for each marginal and the joint distribution in the two cubes. Points are bootstrap-mean estimates and bars are 95% confidence intervals for the inferred cube (posterior-mean) and CTGAN.

### Pairwise associations are robustly preserved in the cube

In race x sex x age, the cube’s Cramér’s V fell within the original’s 95% interval for all three pairs, whereas CTGAN fell slightly below it for the sex x age pair; the mean absolute difference in Cramér’s V across all pairs was 0.037 (95% CI, 0.011-0.071) for the cube versus 0.080 (0.039-0.127) for CTGAN. In the race x sex x 7 medication class x graft rejection cube, the cube followed all the original’s pairwise associations closely, with a mean absolute difference across all pairs of 0.026 (0.019-0.035) versus 0.041 (0.032-0.050) for CTGAN.

### Atomic-cell count vectors are more accurately reconstructed in the cube

R^2^ between the atomic-cell count vectors of the original and the cube was higher than with the CTGAN data in both settings (Table 2): 0.37 (95% CI, 0.17-0.53) in race x sex x age versus −0.33 (−0.59 to −0.10) for CTGAN, and 0.81 (0.70-0.88) in the race x sex x 7 medication class x graft rejection cube versus 0.40 (0.21-0.55) for CTGAN. In both cubes, the cube’s interval lay entirely above CTGAN’s; in race x sex x age, CTGAN’s R^2^ was negative, showing that its atomic-cell counts predicted the original worse than the mean count, while the cube’s stayed positive.

**Table 2.** R^2^ of atomic-cell count vectors against true count vector, by cube and CTGAN synthetic data.

|  | race x sex x age | race x sex x 7 med class x rejection |
| --- | --- | --- |
| **Count Inference** | 0.37 (0.17, 0.53) | 0.81 (0.70, 0.88) |
| **CTGAN synthetic data** | -0.33 (-0.59, -0.10) | 0.40 (0.21, 0.55) |

### Descriptive statistics, odds ratios, and predictive models are preserved

Marginal probabilities estimated from the cube agreed closely with the original, with a smaller mean absolute error than CTGAN for every demographic variable: 0.023 (95% CI, 0.000-0.062) versus 0.050 (0.0003-0.103) for P(sex), 0.017 (0.008-0.029) versus 0.048 (0.033-0.061) for P(race), and 0.010 (0.007-0.014) versus 0.024 (0.020-0.028) for P(age). The cumulative distribution of age from the cube visually overlaid well against the original, while CTGAN deviated substantially (Figure 2).

**Figure 2.**
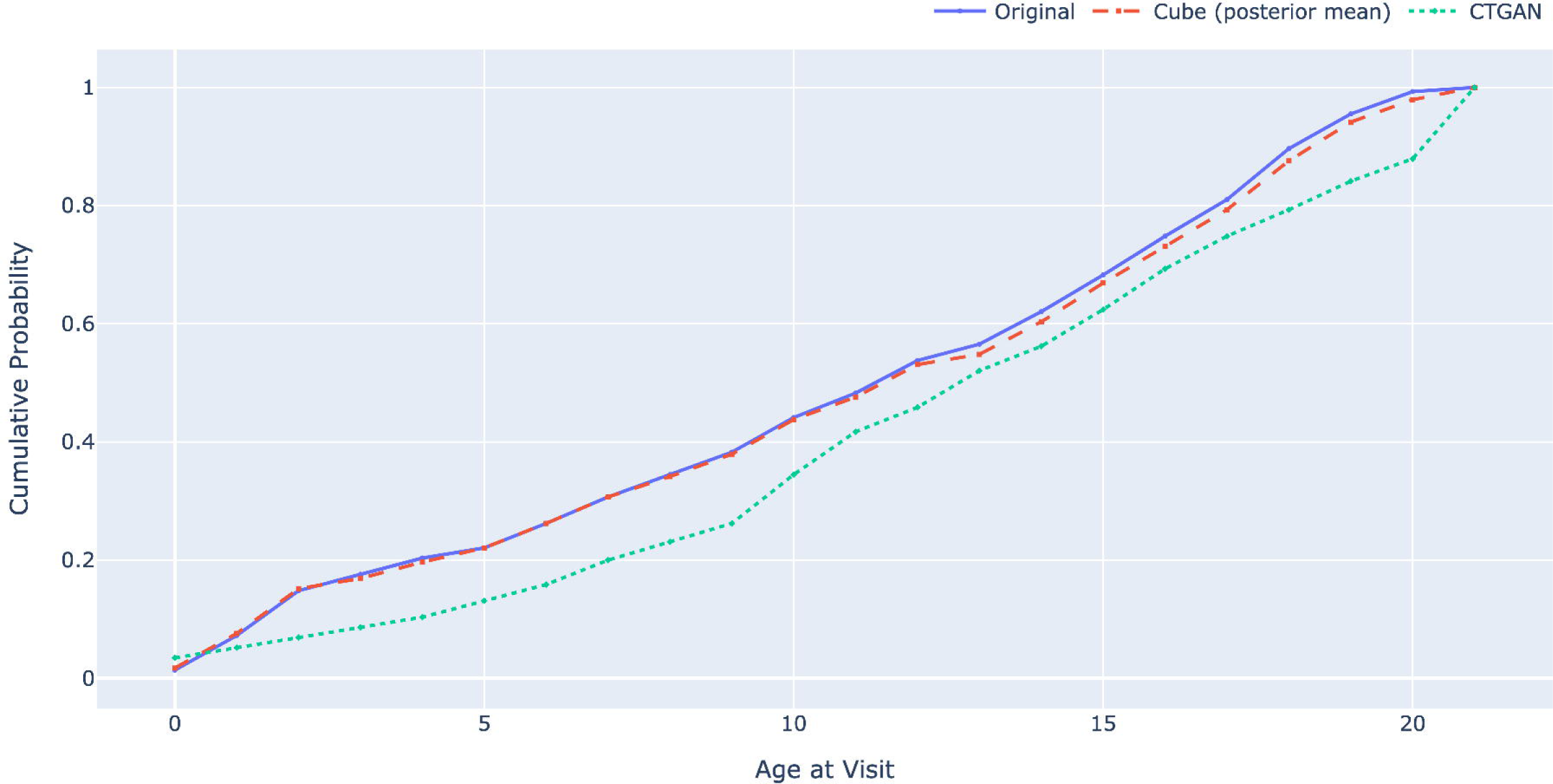
Cumulative distribution of age in the original data, the cube (posterior-mean), and the CTGAN synthetic data.

Across the 106 demographic-by-medication-class subgroups examined for odds ratios of graft rejection, the cube’s log odds ratios departed from the original by a mean absolute error of 1.02 (95% CI, 0.46-1.62) versus CTGAN’s 4.94 (3.82-6.04). The odds ratio direction flipped for a mean of 5.6 subgroups (2-9) under the cube versus 16.1 (12-20) under CTGAN. In the original data, 3.5 subgroups (95% CI, 1-6) showed a statistically significant graft-rejection association. Crucially, none of the cube’s flips involved a significant association (0.0 significant flips; 95% CI, 0-0), whereas a mean of 2.3 of CTGAN’s flips (0-4) reversed an odds ratio significant at p<0.05 in the original data. Because flipped odds ratios would invert clinical interpretation such that a subgroup at apparently elevated risk would appear protected, or vice versa, this finding has direct bearing on the suitability of each surrogate for hypothesis-generating risk analyses.

Logistic-regression classification of graft rejection performed near chance for all three data sources (Table 3), using a 0.5 predicted-probability threshold and showing overlapping test-set bootstrap 95% CIs, reflecting the limited predictive signal carried by the available demographic and medication-class variables in this small cohort. The cube-trained model matched the weak performance of the model trained on the original data, while the CTGAN-trained model was slightly worse on every metric. The comparison is therefore consistent with the cube not introducing spurious predictive signal, rather than demonstrating predictive utility.

**Table 3.** Classification performance for predicting graft rejection.

|  | ORIGINAL | CUBE | CTGAN |
| --- | --- | --- | --- |
| ACCURACY | 0.56 (0.44-0.68) | 0.56 (0.44-0.68) | 0.51 (0.39-0.63) |
| PRECISION | 0.15 (0.03-0.30) | 0.15 (0.03-0.30) | 0.13 (0.03-0.27) |
| RECALL | 0.67 (0.20-1.00) | 0.66 (0.20-1.00) | 0.65 (0.20-1.00) |
| F1 SCORE | 0.24 (0.06-0.43) | 0.24 (0.06-0.43) | 0.22 (0.06-0.40) |

## Discussion

When privacy rules prevent sharing line-level records, this study shows that in this cohort, the cube supported the same descriptive, inferential, and predictive analyses once its suppressed cells were inferred, with conclusions closely resembling those on the original line-level data.

However, the same inference that made it faithful also recovered most of the suppressed atomic cells, including the majority of single-patient cells. Fidelity and privacy are therefore not independent properties to trade off. They are the same reconstruction seen from two sides, so threshold suppression alone cannot deliver one without surrendering the other.

The cube’s overlapping marginal aggregates impose a system of linear equalities and inequalities on the atomic cells. In the 10-variable cube, this system was sufficiently constrained that ILP uniquely determined a majority of suppressed cells, including most single-patient cells.

Suppression of small counts below a threshold is therefore an incomplete disclosure-limitation method for cube releases in this structure. Reconstructing the atomic-cell table is equivalent to recovering the line-level data for these categorical variables, so the same operation that lets analyses recapitulate line-level results also recovers the suppressed small subgroups. The disclosure we demonstrated is a potential existence and attribute disclosure, contingent on auxiliary information — confirming that a small subgroup exists and recovering its suppressed count — rather than re-identification of named individuals, which we did not attempt; even so, for rare attribute combinations, confirming that a single patient occupies a cell can itself be sensitive. Differential privacy offers a formal guarantee for tabular releases by adding calibrated noise to each aggregate count so that any single patient’s contribution is bounded by a privacy-loss parameter, independent of adversary knowledge.[20, 21] A mechanism suitable for a full power-set release would need to account for the entire query workload and the composition of the privacy budget across aggregates, not merely add independent noise to each count. Evaluating how such noise trades off against the fidelity and utility reported here is a natural direction for future work.

The cube’s fidelity was governed by how tightly its published aggregates constrained the suppressed cells. We hypothesize the constraints bind more tightly when the published aggregates are many, overlapping, and each spans fewer cells, conditions that higher dimensionality and lower cardinality generally favor, since more variables yield more overlapping margins and low-cardinality variables split each margin into fewer cells. However, how much is recovered also depends on the data, since the published totals set how tightly each cell is bounded. Dimensionality, cardinality, and cell sparsity vary together across the two cubes and jointly influence recovery. In the cube over race x sex x age, the high-cardinality age variable made most atomic cells sparse and suppressed, with few published counts to anchor inference, so count recovery was more uncertain; even so, the cube preserved pairwise associations at least as well as CTGAN. In the denser 10-variable cube, the overlapping published equations left less uncertainty, with accurate atomic-cell reconstruction and closely preserved Cramér’s V.

Aggregated count releases of this kind are not hypothetical. The SMART/HL7 FHIR Bulk Data Access API[22] provides standardized patient-level extraction at scale, and platforms such as Cumulus[8] use that extraction to assemble cube-style aggregates across participating institutions, with current deployments suppressing counts below k = 10. Our findings have direct bearing on how those releases should be paired with stronger privacy mechanisms. For practitioners weighing this balance, the recommendation is to use the cube for the analytic value it demonstrably carries, but to govern it like line-level data, under a data-use agreement or a formal disclosure-limitation mechanism.

### Limitations

The evaluation used a single, relatively small cohort of pediatric kidney-transplant patients from Boston Children’s Hospital, a single clinical use case; generalizability of these findings to larger cohorts, other disease domains, and other institutions remains to be investigated. The study also examined only categorical variables. Cumulus discretized continuous variables for the power-set aggregation, introducing discretization loss that was not evaluated here. The discretization strategy can itself influence fidelity and utility, an effect left to future work.

The analyses are descriptive and associational rather than prognostic or causal. Each patient’s full longitudinal history was used, immunosuppressive exposures were encoded as ever-exposed indicators, and graft rejection was a single binary outcome, so exposures and outcomes were not placed in a defined temporal order.

For the fidelity and utility evaluations, this study used a single comparator, a synthetic data generation method; the reconstruction evaluation additionally benchmarked count inference against simple count-imputation baselines. While CTGAN is a widely used benchmark for tabular synthesis, a broader comparison with other synthetic data generation methods[16] or privacy-preserving approaches such as differential privacy would provide a more comprehensive interpretation for the cube’s relative performance. We note that this cohort is at the lower end of practical GAN training, so CTGAN performance here should be interpreted as a conservative lower bound for synthetic-data approaches rather than a tight benchmark.

The utility evaluation covered only a small set of downstream analyses relevant to kidney-transplant research. Time-series analyses were outside scope but are a natural next step for more complex tasks. Cohort size itself is limiting, since with so few patients spread across many atomic cells, more complex or high-dimensional multivariable models would have little data to draw on.

The cube reconstruction also faces computational limits as the number of variables grows. The released power set contains a count for every subset of grouping variables, so its size grows exponentially with the number of variables, and the number of atomic cells grows with the product of the variable cardinalities; the reconstruction inherits this scaling, because the integer-programming bounds and the sampler operate over the full set of atomic cells.

## Conclusion

In a single-site cohort of pediatric patients after kidney transplant, aggregated cube data supported descriptive, inferential, and predictive analyses with conclusions consistent with line-level EHR. The same power set structure that enabled this also recovered most of the small cells that suppression was meant to hide. We demonstrated reconstructability of suppressed counts, not re-identification of individuals. Aggregated patient-count releases of this kind should be designed with privacy mechanisms beyond threshold suppression; formal disclosure-limitation methods such as differential privacy warrant evaluation before such releases are scaled to multi-site federated networks.

## Data and Code Availability

The deidentified aggregate counts (cubes) and the analysis code (Bayesian count-inference pipeline, ILP routines, fidelity and utility analyses) used in this study are available from the corresponding author on reasonable request, subject to institutional data-use agreements (DUA). Line-level patient records cannot be shared under the governing IRB approval. A code repository link will be provided at the time of publication.

## Use of AI tools

ChatGPT 5, Claude Opus 4.7, and Grammarly were used to proofread and refine for clarity and readability.

## Supporting information

Supplementary Table S1. Immunosuppressant drug value set

## Data Availability

Fidelity & utility evaluation code are available under Apache License 2.0

https://github.com/smart-on-fhir/cube-inference-fidelity-evaluation

