## Supplementary Table S1. Immunosuppressant drug value set for "Sharing Aggregated Patient Counts in Place of Line-Level EHR Data: Analytic Fidelity and the Limits of Count Suppression for Privacy"

### Supplementary Materials

Supplementary Table S1. Immunosuppressant drug value set

| Drug Class | Ingredient | Abbreviation | Synonyms |
| --- | --- | --- | --- |
| Anti-metabolite | Azathioprine | AZA | - Azathioprine - Imuran - Azasan - Thiopurine immunosuppressant - 6-Mercaptopurine - 6 Mercaptopurine |
| Anti-metabolite | Mycophenolate Mofetil | MMF | - Mycophenolate - MMF - CellCept - Myfortic - Mycophenolic Acid |
| Calcineurin Inhibitor (CNI) | Cyclosporin | CYA | - Cyclosporin - Cyclosporine - Ciclosporin - Neoral - Sandimmune - Gengraf - Restasis - Cequa |
| Calcineurin Inhibitor (CNI) | Tacrolimus | TAC | - Tacrolimus - FK506 - FR-900506 - Prograf - Advagraf - Fujimycin - Envarsus |
| Corticosteroid | Methylprednisolone | Medrol | - Methylprednisolone - Medrol - Solu-Medrol - Depo-Medrol - A-Methapred |
| Corticosteroid | Prednisolone | PDL | - Prednisolone - Orapred - Pediapred - Prelone - Flo-Pred - Millipred - Veripred |
| Corticosteroid | Prednisone | PRED | - Prednisolone - Orapred - Pediapred - Prelone - Flo-Pred - Millipred - Veripred |
| Costimulation blocker | Belatacept | BEL | - Belatacept - Nulojix |
| mTOR inhibitor | Everolimus | EVE | - Evorolimus - Zortress - Afinitor - Votubia - RAD001 - SDZ-RAD |
| mTOR inhibitor | Sirolimus | SRL | - Sirolimus - AY-22989 - Rapamune - Rapamycin - Streptomyces hygroscopicus - Macrolide immunosuppressant |
| Polyclonal antibody | Antithymocyte Globulin | ATG | - ATG - Anti-Thymocyte Globulin - Anti Thymocyte Globulin |
| Monoclonal  antibody (mAb) | Rituximab | RTX | - Rituxan - Truxima - Ruxience - IDEC-102 - MabThera - Reditux - RTXM 83 |
| Monoclonal  Antibody (mAb) | Alemtuzumab | ALEM | - Alemtuzumab - Campath - Lemtrada - LDP-03 - MabCambath |
| Monoclonal  Antibody (mAb) | Basiliximab | BAS | - Basiliximab - Simulect - CHI 621 |
